# Disorder-and cognitive demand-specific neurofunctional alterations during social emotional working memory in generalized anxiety disorder and major depressive disorder

**DOI:** 10.1101/2022.01.18.22269466

**Authors:** Xiaolei Xu, Fei Xin, Congcong Liu, Yuanshu Chen, Shuxia Yao, Xinqi Zhou, Feng Zhou, Yulan Huang, Jing Dai, Jinyu Wang, Zhili Zou, Keith M Kendrick, Bo Zhou, Benjamin Becker

## Abstract

Generalized Anxiety Disorder (GAD) and Major Depressive Disorder (MDD) are both characterized by cognitive and social impairments. Determining disorder-specific neurobiological alterations in GAD and MDD by means of functional magnetic resonance imaging (fMRI) may promote determination of precise diagnostic markers. This study aimed to examine disorder-specific behavioral and neural alterations at the intersection of social and cognitive processing in treatment-naïve first-episode GAD (n=35) and MDD (n=37) patients compared to healthy controls (n=35) by employing a social-emotional n-back fMRI paradigm. No behavioral differences between patients and healthy controls were observed. However, GAD patients exhibited decreased bilateral dorsomedial prefrontal cortex (dmPFC) engagement during the 0-back condition yet increased dmPFC engagement during the 1-back condition compared to MDD and healthy participants. In contrast, MDD patients exhibited increased dmPFC-insula coupling during 0-back, yet decreased coupling during 1-back, compared to GAD and healthy participants. Dimensional symptom-load analysis confirmed that increased dmPFC-insula connectivity during 0-back was positively associated with depressive symptom load. These findings suggest that the dmPFC engaged in integrating of affective and cognitive components and self-other processing exhibits GAD-specific neurofunctional dysregulations whereas functional dmPF communication with insula, a region involved in salience processing, may represent an MDD-specific neurofunctional deficit.

## Introduction

Depression and anxiety are characterized by highly debilitating impairments in emotional, cognitive and social domains. In the pathological range particularly Major Depressive Disorder (MDD) and Generalized Anxiety Disorder (GAD) exhibit high levels of comorbidity [1-3]. Both disorders are characterized by overlapping emotional dysregulations and cognitive deficits on the symptomatic level, including impaired social processing and executive functions [3, 4-6] which leads to an ongoing debate about common and specific neurobiological markers to distinguish these disorders.

Working memory is a fundamental executive cognitive function allowing individuals to temporarily maintain and manipulate a limited amount of information in different domains for immediate use [7]. Cognitive working memory (CWM) refers to the processing of perceptual or cognitive information such as digits or shapes while social working memory (SWM) refers to social cognitive information such as faces or other people’s mental states with the latter being particular critical for social functioning in daily life [8, 9]. Despite convergent evidence for marked social impairments in both, GAD and MDD, and SWM deficits being proposed as an underlying transdiagnostic candidate mechanism [10] common and separable neural and behavioral dysregulations in this domain have not been examined.

Functional magnetic resonance imaging (fMRI) studies have demonstrated that both CWM and SWM critically depend on the structural and functional integrity of the prefrontal cortex and dynamic interactions between the frontoparietal control and the default mode network (DMN) [11-14]. However, SWM additionally relies on systems involved in social processes such as the mentalizing and face processing networks engaged in determining other people’s intentions and mental states or identity and emotional states, respectively [9, 15-17]. The dorsomedial prefrontal cortex (dmPFC) represents a core node of the mentalizing network and plays an important role in integrating cognitive and affective components of SWM [18]. In addition, on the network level the insula has been increasingly identified as a potential core hub that supports attention deployment towards the salience of social stimuli [19-21] probably via its role in switching between large scale networks engaged in attention orientation such as the central executive network (CEN) and DMN to facilitate access to working memory and attention resources [22, 23].

Working memory refers to a complex cognitive function which requires a continuous trade-off between stability versus flexibility [24]. Particular crucial subprocesses for this balance encompass maintenance referring to the ability to maintain stored information despite irrelevant distractors and updating referring to the ability to continuously update the limited storage capacity according to new information [25, 26]. Specifically, selectively updating goal-relevant information in SWM with emotional content plays an important role in social-emotional and social-cognitive deficits in psychiatric disorders [27], such that previous studies have demonstrated impaired SWM maintenance and updating in both, patients with anxiety and depression [27-29].

Several neuroimaging case control studies employed fMRI to compare either GAD or MDD patients with healthy controls revealed that both disorders exhibited neurofunctional abnormalities during both CWM and SWM processing [4, 30]. Compared to healthy subjects, patients with GAD exhibited decreased gray and white matter volumes as well as functional engagement in the dorsolateral prefrontal cortex (dlPFC) specifically during SWM when negative emotional distractors were involved [31-34]. While GAD has been characterized by prefrontal hypoactivation, hyperactivation in these regions particularly the left dlPFC has been consistently found in MDD patients [35, 36].

Previous meta-analysis of the case control neuroimaging studies in MDD demonstrated widespread functional and structural cortical-limbic-subcortical dysfunctions encompassing regions such as dlPFC, insula as well as temporal and parietal regions which may underly working memory as well as social deficits in MDD [30, 33].

Although a large number of case control studies examined the neural alterations in GAD and MDD separately, these studies are limited with respect to determining common and disorder-specific neurobiological dysregulations [37]. An increasing number of studies thus employed transdiagnostic designs including both, GAD and MDD patients and were able to determine disorder-specific insula alterations during pain empathy [38], lateral prefrontal alterations during emotional context-specific cognitive control [39, 40], and network level alterations in frontal executive control networks [41, 42]. Given the overlap of these regions with social processing and working memory core regions we aimed at determining whether GAD and MDD exhibit separable neurofunctional dysregulations during SWM.

Against the background, the present study aims to examine common and distinct behavioral and neural alterations during SWM in unmedicated first episode GAD and MDD by employing a social n-back task [12, 43]. The n-back paradigm has been widely validated to examine CWM and SWM and has a high specificity to both maintenance and updating. Briefly, during this paradigm a stream of stimuli is presented, and participants are required to indicate whether the current stimulus matches the one which is presented on n trials before [44]. We capitalized on a previous version which employed emotional facial expressions as stimuli and participants were asked to focus on either the identity (ID) or the emotion (EMO) of the same set of facial stimuli [43]. This emotional n-back version allows us to investigate different aspects of SWM content updating such as identity and emotional expression [45, 46]. In addition, the control 0-back task is normally used to investigate the maintenance component and when n>0 the n-back task draws upon both maintenance and updating processes as every stimulus has to be maintained and further serves as updated target for the subsequent trial [13].

fMRI studies have demonstrated that the key regions and networks engaged in SWM processes (e.g. frontoparietal system, default network) can be successfully activated by employing the emotional n-back paradigm [44, 47, 48]. Given that previous case-control studies revealed opposing prefrontal dysregulations in GAD and MDD during n-back paradigms [35, 49, 50] and the critical role of prefrontal regions in both, working memory and social processing we hypothesized disorder-specific neurofunctional dysregulations in these regions.

## Methods

### Participants

72 treatment-naïve first-episode MDD (n = 37) and GAD (n = 35) patients were recruited via the Sichuan Provincial People’s Hospital and The Fourth People’s Hospital of Chengdu (Chengdu, China). Healthy controls (HC, n = 35) were recruited by local advertisements (Exclusion criteria for patients and HC see Supplementary materials). GAD and MDD diagnosis were determined by an experienced psychiatrist according to DSM-IV (Sichuan Provincial People’s Hospital) or ICD-10 (The Fourth People’s Hospital of Chengdu) criteria and further confirmed by an experienced psychologist using Mini International Neuropsychiatric Interview (M.I.N.I.) for DSM-IV [51]. As part of a large project examining the common and disorder-specific alterations in GAD and MDD, participants had completed a resting state fMRI acquisition [42], a pain empathy paradigm [38] and a go/no-go task [40] sequentially before further completing this task. Depression and anxiety symptom load were assessed by the Beck Depression Inventory II (BDI-II, [52]) and Penn State Worry Questionnaire (PSWQ, [53]) respectively. All participants provided written informed consent after they were informed about the detailed study procedures and informed that they were allowed to withdraw from this study at any time without negative consequences. This study and all procedures were fully approved by the local UESTC ethics committee and adhered to the latest revision of the Declaration of Helsinki.

### Working memory paradigm

A previously validated social-emotional n-back paradigm [43] employing facial stimuli was employed in the present study with two levels of cognitive load: 0-back and 1-back. The paradigm included two face recognition conditions: (1) facial emotion expression (EMO) recognition, or (2) facial identity (ID) recognition, leading to a total of four working memory load times social processing conditions: 0-back targeting emotion (0EMO), 0-back targeting identity (0ID), 1-back targeting emotion (1EMO), and 1-back targeting identity (1ID). During the 0-back condition participants were asked to indicate whether the present face matched with a designated face in terms of identity or emotional expression presented at the beginning of the respective task block. During the 1-back condition participants were asked to indicate whether the current stimulus was identical to the previous one in terms of identify or emotional expression respectively. A total of four runs including four randomized blocks (one block per condition) was used in the present study and 16 faces from four actors (two females and two males) posing four expressions (neutral, happy, angry, and fearful) from the facial database in our lab were randomly presented within each block [54, 55]. Within each block, each facial stimulus was displayed for 2000ms followed by a 500ms fixation cross (**Figure 1)**. Each block started with a 4s instruction indicating the task requirement for the next block and blocks were interspersed by 16s low-level inter-block-intervals.

**Fig 1.**
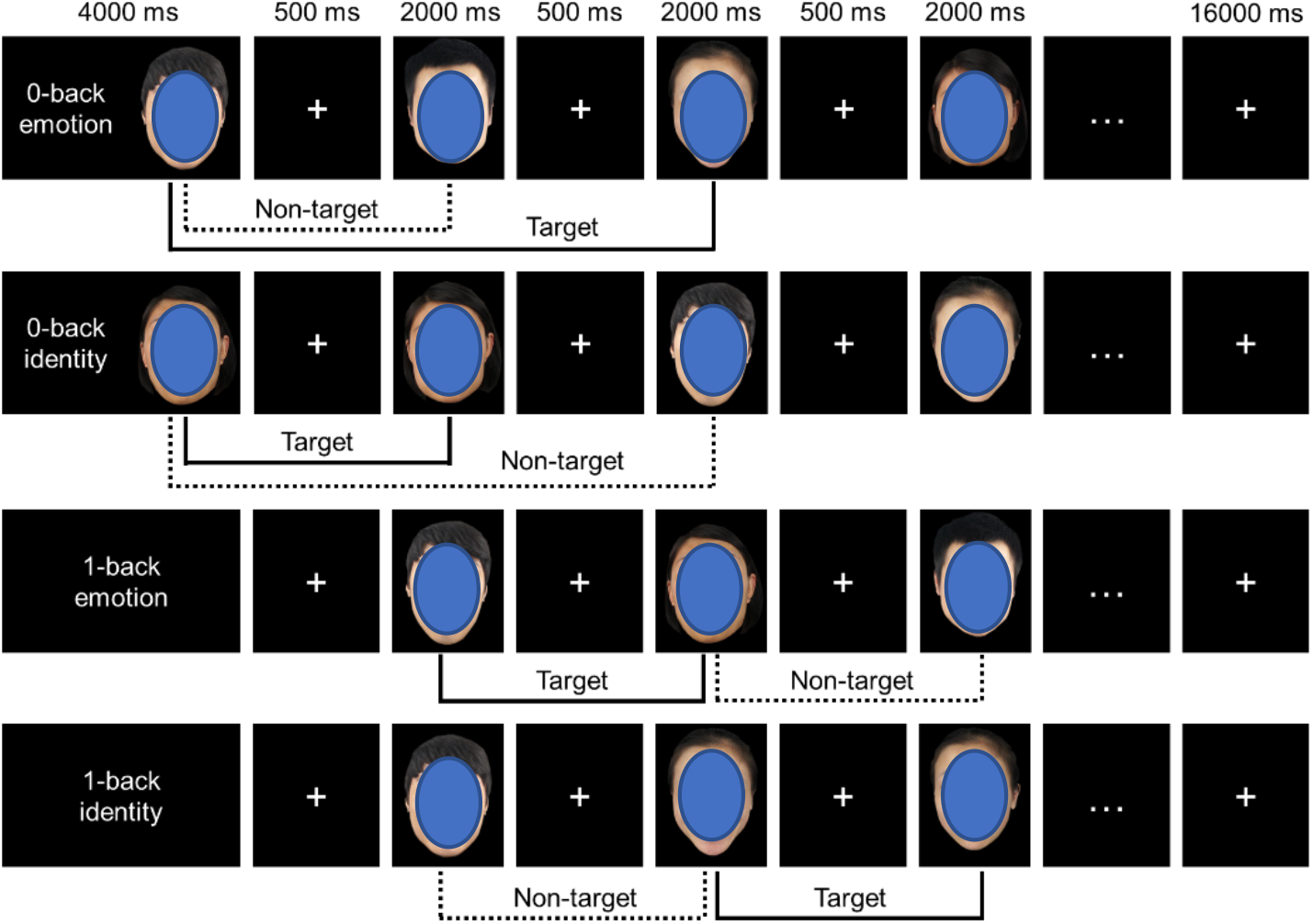
The emotional working memory task. In the 0-back condition participants were required to judge whether the current stimulus was identical to a designated stimulus presented at the beginning of the block. In 1-back condition participants were required to judge whether the current stimulus matches the previous one. Depending on the instruction provided at the beginning of the block subjects had to focus on the emotion or identity of the faces, respectively. Note: for the preprint version oval shapes were used to cover the example faces.

### MRI data acquisition

MRI data were collected using a 3.0 Tesla GE MR750 system (General Electric Medical System, Milwaukee, WI, USA). Scanning parameters are reported in Supplementary materials.

### MRI data preprocessing

MRI data was processed and analyzed using SPM12 (Welcome Trust Center for Neuroimaging, University College London, London, United Kingdom). To allow magnet-steady images the first 6 functional volumes were discarded. Preprocessing for the remaining functional images included slice timing, realigning to correct for head motion, normalization into Montreal Neurological Institute (MNI) employing a two-step procedure implementing the segmentation of the T1 images and application of the resulting transformation parameters to the functional time series resampled at 3mm^3^ voxel size and spatial smoothing using an 8mm full-width at half-maximum (FWHM) Gaussian kernel.

A two-level random effects general linear model (GLM) analysis was employed to the fMRI data for statistical analyses [56]. On the first level the four experimental conditions were modelled using a box-car function (‘0EMO’, ‘0ID’, ‘1EMO’, ‘1ID’) and convolved with the canonical hemodynamic response function (HRF). To further control motion-related artifacts the six head-motion parameters were included in the design matrix. The inter-block interval served as implicit baseline. In line with previous studies the main interaction contrast of interest [(1EMO > 1ID) > (0EMO > 0ID)] and emotion-load contrasts [1EMO > 1ID]; [0EMO > 0ID] were subjected to second level group-level analysis (see e.g. [43]). Common and separable neural alterations between GAD and MDD were examined by means of a voxel-wise whole-brain mixed ANOVA in SPM with the contrast [(1EMO > 1ID) > (0EMO > 0ID)] and group as between-subject factor (corrected for multiple comparison using FWE cluster-level correction and *p* <.05, initial cluster forming threshold *p* <.001, as recommended by Woo et al., 2014 [57] and Slotnick, 2017 [58].

To further determine disorder-specific alterations on the network level a generalized form of context-dependent psychophysiological interaction analysis (gPPI) [59] was conducted to examine task-dependent functional connectivity using a seed-to-voxel approach on the whole brain level. Seed regions were determined based on the findings from the neural reactivity changes. Modelling and conditions on the first level were identical to the BOLD activation analyses and the identical main contrast of interest [(1EMO > 1ID) > (0EMO > 0ID)] was subjected to the voxel-wise whole brain second level ANOVA analysis.

In addition to this categorical approach employing the diagnostic groups, a dimensional analysis was employed to explore associations between neural indices and depressive (BDI-II scores) and GAD (PSWQ scores) symptom-load in the entire sample [60] (details see Supplementary materials).

## Results

### Demography and dimensional symptom load

According to the exclusion criteria 26 subjects were excluded leading to a final sample of n=81 (HC=29, GAD=25, MDD=27, details see Supplementary materials). Participants in GAD, MDD and HC groups were matched for age (*p*=0.219) and education level (*p*=0.824). Chi-square test for gender revealed significant gender difference between groups indicating a higher proportion of women in the MDD group than GAD and HC groups (*p*_(X)_ ^2^ = 0.003, details see **Table 1**). Univariate ANOVA was employed to examine group differences in symptom load and early life stress. Results revealed significant main effects of group for GAD symptom load (F_2, 71_ = 54.762, *p*<0.001, η^2^ = 0.607), MDD symptom load (F_2, 71_ = 65.348, *p*<0.001, η^2^ = 0.648) and early life stress (F_2, 71_ = 4.661, *p*<0.05, η^2^ = 0.116). Post-hoc analyses indicated that GAD symptom load was higher in both GAD and MDD disorders compared to HC, but not significantly different between the two disorders. Depressive symptom load was higher in both patient groups compared to HC, and in MDD compared to GAD patients (marginal significant, *p* = 0.053). In addition, MDD patients reported significantly higher early life stress compared to HC whereas no differences were found between GAD and HC (see table 1 for details).

**Table 1.**
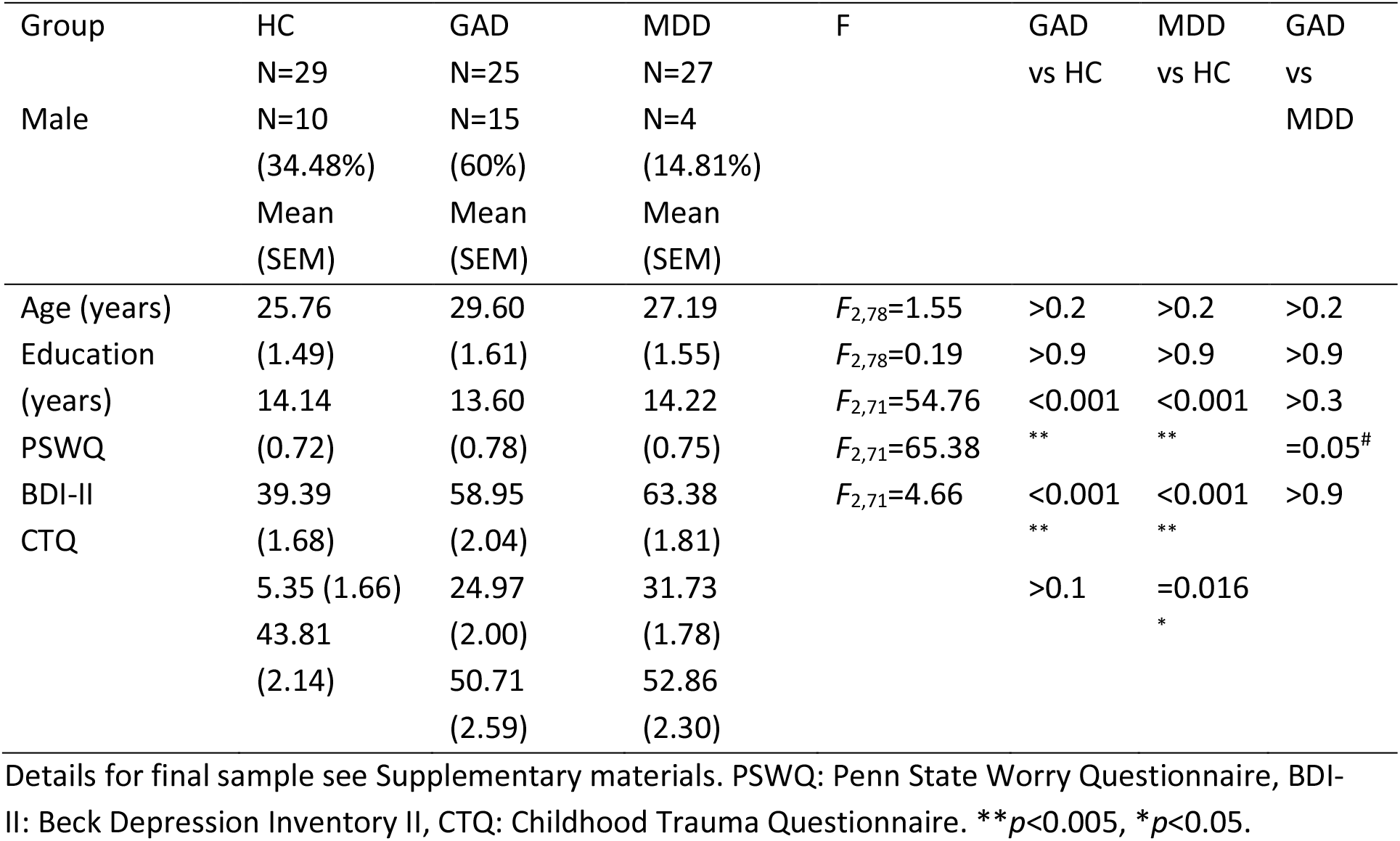
Demographics, symptom load and early life stress (mean, SEM)

### Behavioral results

The mixed ANOVA load (0-back/1-back) x social (emotion/identity) x disorder (GAD/MDD/HC) with gender as covariate was employed to examine working memory performance in terms of accuracy and response time. Examining accuracy revealed a significant main effect of social (F1,77 = 5.276, *p* = 0.024, η_p_^2^= 0.064), and interaction effects in terms of load x social (F1,77 = 5.439, *p* = 0.022, η_p_^2^ = 0.066) and load x social x disorder (F1,77 = 3.702, *p* = 0.029, η_p_^2^ = 0.088). Post-hoc Bonferroni-corrected paired comparison showed that the accuracy for identity was significantly higher than for emotion (emotion=89.7%±0.01, identity=93.3%±0.01) and interactions were mainly driven by higher accuracy identity recognition across both working memory load conditions (details see Supplementary materials). Regarding response times, results showed a significant main effect of social (F1,77 = 48.474, *p* < 0.001, η_p_^2^ = 0.386) and a marginal significant main effect of load (F1,77 = 3.904, *p* = 0.052, η_p_^2^ = 0.048) with post-hoc Bonferroni-corrected paired comparison indicating that participants responded faster at the low working memory load (0-back = 657.079ms±11.955, 1-back = 757.583ms±12.532, *p*<0.001) and identity (identity = 648.015ms±12.077, emotion = 766.646±12.132, *p*< 0.001) compared to the higher load or emotion recognition, respectively. Overall, no between group significant differences were observed.

### Neuroimaging results

#### Social working memory activation

Examining disorder-specific differences in neural activity as a function of social processes and working memory load by means of a voxel-wise mixed ANOVA including disorder group as between-subject factor and gender as covariate revealed a main effect of group in the bilateral dorsomedial prefrontal cortex (dmPFC, MNI: left dmPFC [0 57 21], right dmPFC [6 54 36], k = 228, *p*_*F*WE-cluster_ = 0.001, **Fig.2a**). Post-hoc comparison by means of subjecting extracted parameter estimates from this region to an SPSS one-way ANOVA with group as between-subject factor indicated that GAD patients exhibited increased dmPFC activity compared to both MDD patients and HC. No significant alteration was found in MDD patients compared to HC. Further disentangling the interaction effect revealed disorder- and social processing-specific neurofunctional dmPFC dysregulations such that GAD patients exhibited decreased reactivity at the low working memory load yet increased reactivity during the higher working memory load for emotion versus identity recognition as compared to both MDD and HC (**Fig. 2b**).

**Figure 2.**
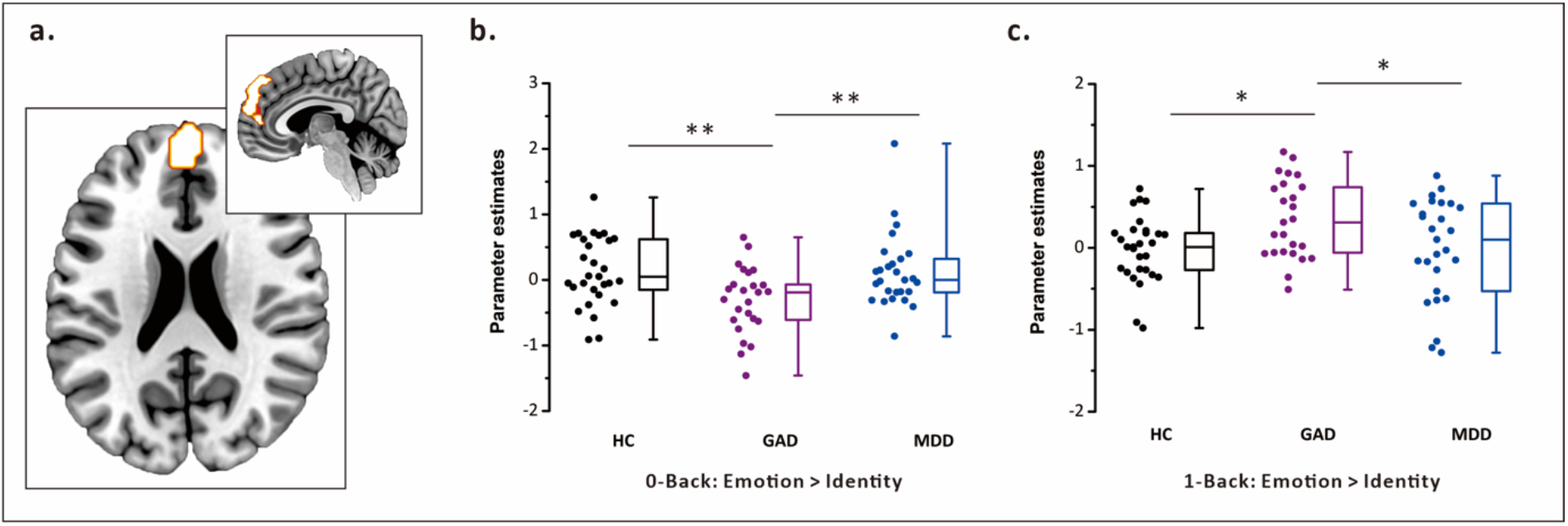
**a**. Voxel-wise whole-brain ANOVA using cluster level FWE correction revealed a significant interaction effect in the bilateral dorsomedial prefrontal cortex (dmPFC). **b**. Activation in the dmPFC was decreased during 0-back and **c**. increased during 1-back working memory load in GAD patients compared to both healthy control (HC) and patients with major depressive disorder (MDD). \**p*<0.05, \*\**p*<0.01.

#### Network analysis

To explore alterations on the network level, a gPPI analysis was performed using the identified dmPFC cluster as seed and the contrast of interest ([1EMO > 1ID]>[0EMO > 0ID]) was subjected to the voxel-wise whole brain group level comparison. Results revealed an interaction effect in the left dorsal anterior insula (dAI, MNI: [-36 -3 15], k = 51, *p*_FWE_ = 0.021, **Fig. 3a**). Post-hoc comparison indicated that connectivity between dmPFC and insula was decreased in MDD patients compared to both GAD patients and HC. No significant alteration was found in GAD patients compared to HC. Moreover, disentangling the interaction revealed that MDD patients exhibited increased dmPFC-dAI connectivity during 0-back compared to HC group whereas decreased connectivity during 1-back compared to both HC and GAD group (**Fig. 3b**).

**Figure 3.**
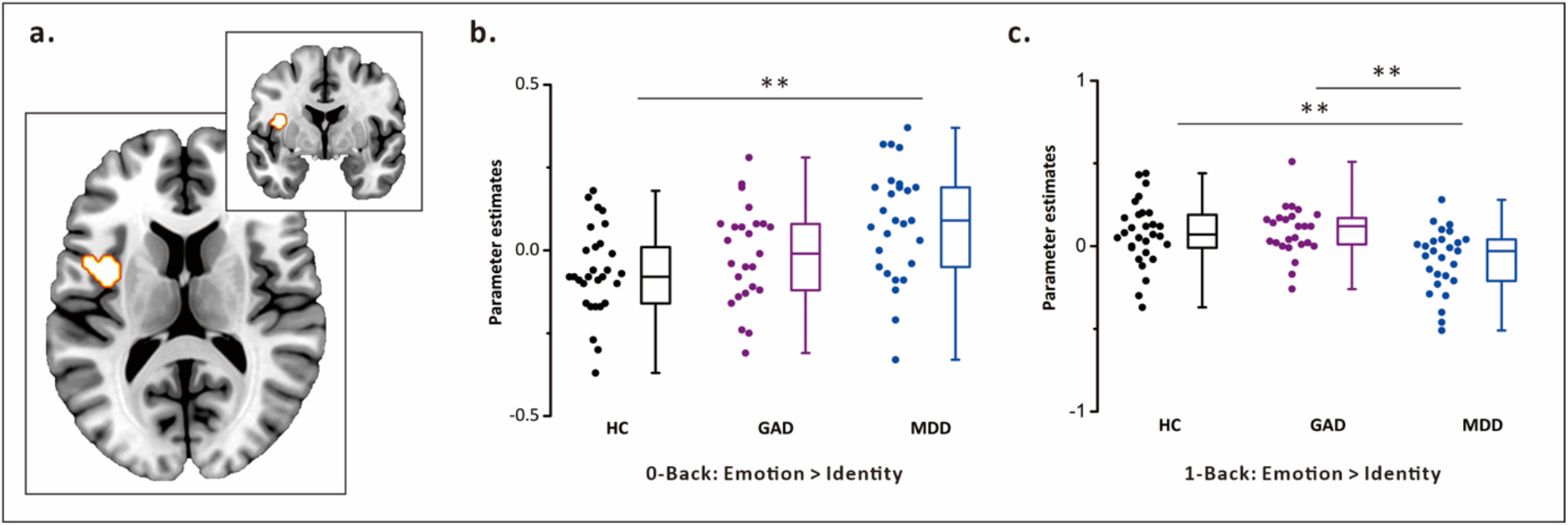
**a**. Voxel-wise seed-to-whole brain connectivity analysis examining dmPFC functional connectivity differences between the groups revealed a significant group times social times working memory load interaction effect on the functional coupling between the dorsomedial prefrontal cortex (dmPFC) and the dorsal anterior insula (dAI) such that coupling in this pathway was **b**. increased during 0-back in MDD compared to HC and **c**. decreased during 1-back in MDD compared to both HC and GAD patients. \*\**p*<0.01.

#### Dimensional analyses in the entire sample: associations between neural indices and symptom load

Results from dimensional analyses indicated that depressive symptom load was positively associated with dmPFC-dAI connectivity (r = 0.27, *p*_*FWE*_ = 0.036, **Fig. 4**) while controlling for GAD symptom load. No other associations were found between neural indices and symptom load.

**Figure 4.**
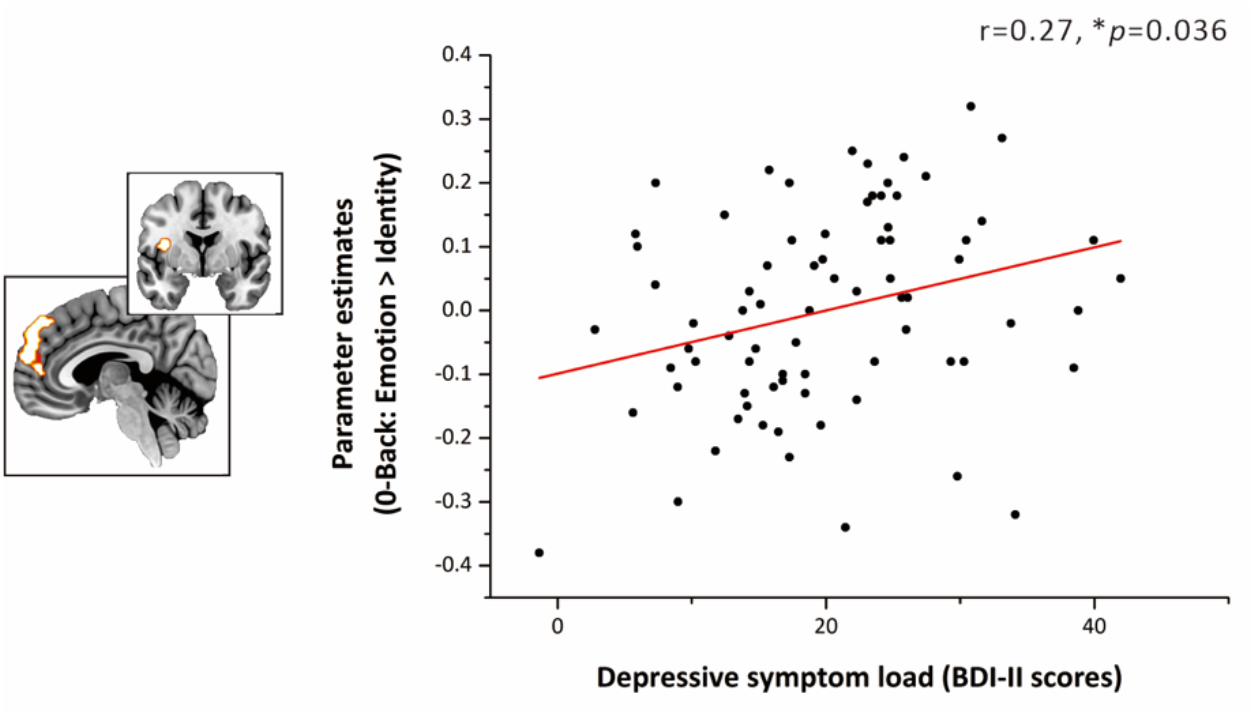
Association between dmPFC-dAI connectivity with depressive symptom load in the entire sample. \**p*<0.05.

## Discussion

This study employed a social n-back fMRI paradigm to determine common and disorder-specific neurofunctional alterations during SWM processing by means of both categorical and dimensional analyses in treatment-naïve unmedicated GAD and MDD patients. No behavioral differences were found between patient groups and healthy control. On the neural level the dmPFC reactivity was reduced during 0-back and increased during 1-back cognitive load specifically in GAD patients compared to both healthy controls and MDD patients. However, network analysis revealed that the identified dmPFC - dAI connectivity was increased during 0-back and decreased during 1-back specifically in MDD patients compared to healthy controls. Dimensional analysis further confirmed that the severity of depression reflected by the BDI score was positively associated with the dmPFC – dAI coupling during the 0-back condition in the entire sample.

Consistent with previous findings participants in the present study showed higher accuracy and shorter response times to identity than emotion during both 0- and 1-back working memory load which may suggest that emotion recognition requires more cognitive resources than identity recognition [43]. However, no significant group differences were found suggesting similar performance between GAD, MDD patients and healthy subjects. Our findings resemble previous findings showing that both GAD and MDD patients performed worse than healthy control during high (e.g. 2- and 3-back) but not low (e.g. 0-back) working memory conditions [61, 62].

On the neural level, altered dmPFC activation may represent a GAD-specific dysfunction in the context of maintaining and updating social emotion vs. identity information. The dmPFC is a key region in social cognition including emotional memory, self-referential processing and decision-making [63, 64]. This region integrates cognitive and affective components for further processing stages and integration with memory while additionally emphasizing or de-emphasizing the impact of emotional content on maintenance, storage and retrieval of information [18]. In the domain of social processing the dmPFC plays an important role in mentalizing and learning to decode affective states of others while integrating this information with self-referential information in memory [65, 66]. Dysregulated dmPFC activity in GAD patients during social emotional recognition working memory may reflect an impaired ability to decode the affective states of others and while balancing the emotional and cognitive requirements to optimize task performance in social contexts. In addition, GAD patients may fail to learn from and utilize previous experiences to facilitate proper decision-making in social interactions in turn promoting insecurity and rumination.

In contrast, on the network level individuals with MDD exhibited increased dmPFC-dAI communication during 0-back and decreased communication between them during 1-back condition compared to both, GAD and healthy controls. As a key node in the salience network (SN) the anterior insula plays a prominent role in detecting salient events via bottom-up attention processing and facilitates the integration of attention and working memory by switching between SN and other large-scale networks including the CEN and DMN [67-69]. Several previous studies reported dysregulated network communication in these networks in MDD [70-72]. Altered communication of the dmPFC - key social and self-referential processing node – with the anterior insula may thus suggest that MDD patients augment the salience of self-centered rumination during daily social interactions therefore leading to a sustained depressive state. Evidence from recent Transcranial Magnetic Stimulation (TMS) experiments demonstrated that targeting the dmPFC can reduce depression symptoms by decreasing the connectivity between dmPFC and insula [73]. Together with a previous study reporting that intrinsic connectivity of the prefrontal-insular network may represent an MDD-specific signature [74] our findings indicate that dmPFC-dAI connectivity alteration may represent an MDD-specific neural marker during social emotional working memory processesing. In addition, an exploratory dimensional analysis indicated that dmPFC-dAI communication was associated with the level of depressive symptoms in the entire sample, resembling previous findings on a disorder-specific involvement of these pathways in depression [74].

Notably, dmPFC activation in GAD patients was decreased during 0-back but increased during 1-back while dmPFC-dAI connectivity in MDD patients was increased during 0-back while decreased during 1-back cognitive load. The opposing neural alterations during the 0- and 1-back conditions may indicate different neural alterations during maintenance and updating processing in SWM. Higher cognitive 1-back working memory relies on dynamic maintaining and updating of new stimuli therefore the interaction between emotional and cognitive processing may be more complex than 0- back condition which only includes maintenance of a priori designated stimuli.

There are several limitations in the present study. First, our strict enrollment criteria led to a moderate sample size. Second, this sample size did not allow us to further explore gender differences although male and female patients with GAD and MDD have been demonstrated to show significant difference in neural activity [75, 76]. Third, according to the M.I.N.I. interview some patients (MDD = 7 in the GAD group, GAD = 6 in the MDD group) exhibited secondary MDD or GAD comorbidity although the primary diagnosis of MDD and GAD was determined by experienced clinical psychiatrists. Finally, the emotional working memory paradigm and the block design we employed in the present study did not allow us to further examine the specific emotional impacts to working memory.

Together, findings from the present study suggest that the prefrontal regions specifically the dmPFC which is highly engaged in integrating affective and cognitive components of memory represents disorder-specific neurofunctional alteration during emotional working memory for GAD. However, the deficient prefronto-insular communication which is engaged in facilitating the integration of salience attention and memory by switching between different networks may represent the neurofunctional biomarker for MDD.

## Data Availability

All data produced in the present study are available upon reasonable request to the authors

## Funding and disclosure

This work was supported by the National Key Research and Development Program of China (2018YFA0701400), and the National Natural Science Foundation of China (NSFC, 61871420, 32100887), and China Postdoctoral Science Foundation (2020M673165). The authors declare no competing interests.

## Author contributions

XX, BB, FX, and KMK designed the experiment. XX and FX prepared the study protocols and procedures. BZ, JD, ZZ, YH, and JW performed the clinical assessments. XX, FX, YC and CL acquired data. XX, FX, XZ, FZ and SY analyzed the data. XX and BB interpreted the data and drafted the paper. All authors commented on and gave final approval to the final version of the paper.

## Supplementary Materials

### Exclusion criteria, initial quality assessments and final sample

All patients were treatment-naïve and had not previously received a diagnosis of or treatment for any psychiatric disorders. The diagnostic assessments were conducted during initial admission to the hospitals by experienced psychiatrists and suitable patients underwent the fMRI assessments during the period of further diagnostic clarification without receiving any treatment (<5 days after admission). The following exclusion criteria were applied to all participants, including controls: (1) history of or current episode of the following axis I disorders according to DSM criteria: post-traumatic stress disorder, PTSD, feeding and eating disorders, substance use disorders, bipolar disorder, and mania, (2) history of or current clinically relevant medical or neurological disorder, (3) acute (within six weeks before the assessments) or chronic use of medication, (4) acute suicidal tendencies, (5) contraindications for MRI assessments, (6) left handedness, (7) incapable of understanding the task, and, (8) technical issues or excessive motion during MRI assessment (head motion >3mm).

All healthy controls were without psychiatric disorders according to the M.I.N.I. interview. Several patients (and one HC) were too exhausted to continue with the questionnaires following the MRI assessments leading to the number of participants per group varying from 23 to 29 (BDI-II, PSWQ) and 21-28 (CTQ) respectively. Importantly, there was no significant difference in the number of participants remaining for analyses between groups (χ^2^ = 0.296, *p* = 0.862).

Study exclusion criteria, inspection of data quality and the independent diagnostic interview led to exclusion of n = 26 subjects due to technical issues during fMRI data collection (n=3), unable to follow task instructions (n=12), head motion > 3mm (n=2), MDD and GAD diagnosis not validated by the M.I.N.I. or presence of a co-morbid previous or current disorder in the M.I.N.I. according to the exclusion criteria: PTSD (n=2), OCD (n = 1) (as primary diagnosis, GAD not confirmed by the M.I.N.I.), substance use disorder (n = 1), bulimia nervosa (n = 1), agoraphobia (n = 1), (as primary diagnosis, MDD diagnosis not confirmed in the M.I.N.I.), acute suicidal tendencies (n = 1), and mania (n = 2). The final sample for all fMRI analyses included HC = 29, GAD = 25, and MDD = 27 (total n = 81, details see **Figure S1**). All patients in the GAD group received the primary diagnosis GAD, all patients in the MDD group received the primary diagnosis MDD according to DSM criteria across the diagnostic assessment. Given the high prevalence of unipolar depressive and anxiety-associated disorders in GAD or MDD, respectively, a secondary diagnosis in these disorders that was additionally determined by the M.I.N.I. interview was thus not considered as exclusion criterion. N = 13 patients in the GAD (total n = 25) group and n = 14 (total n = 27) patients in the MDD group did not exhibit an additional psychiatric diagnosis. The following secondary diagnoses were determined by the M.I.N.I. interview: social phobia (GAD, n = 3, MDD, n = 3), obsessive compulsive disorder (GAD, n = 2; MDD, n = 2), panic disorder (GAD, n = 5; MDD, n = 1), agoraphobia (GAD, n = 5; MDD, n = 4), MDD (n = 7 in the GAD group), GAD (n = 6 in the MDD group).

**Figure S1.**
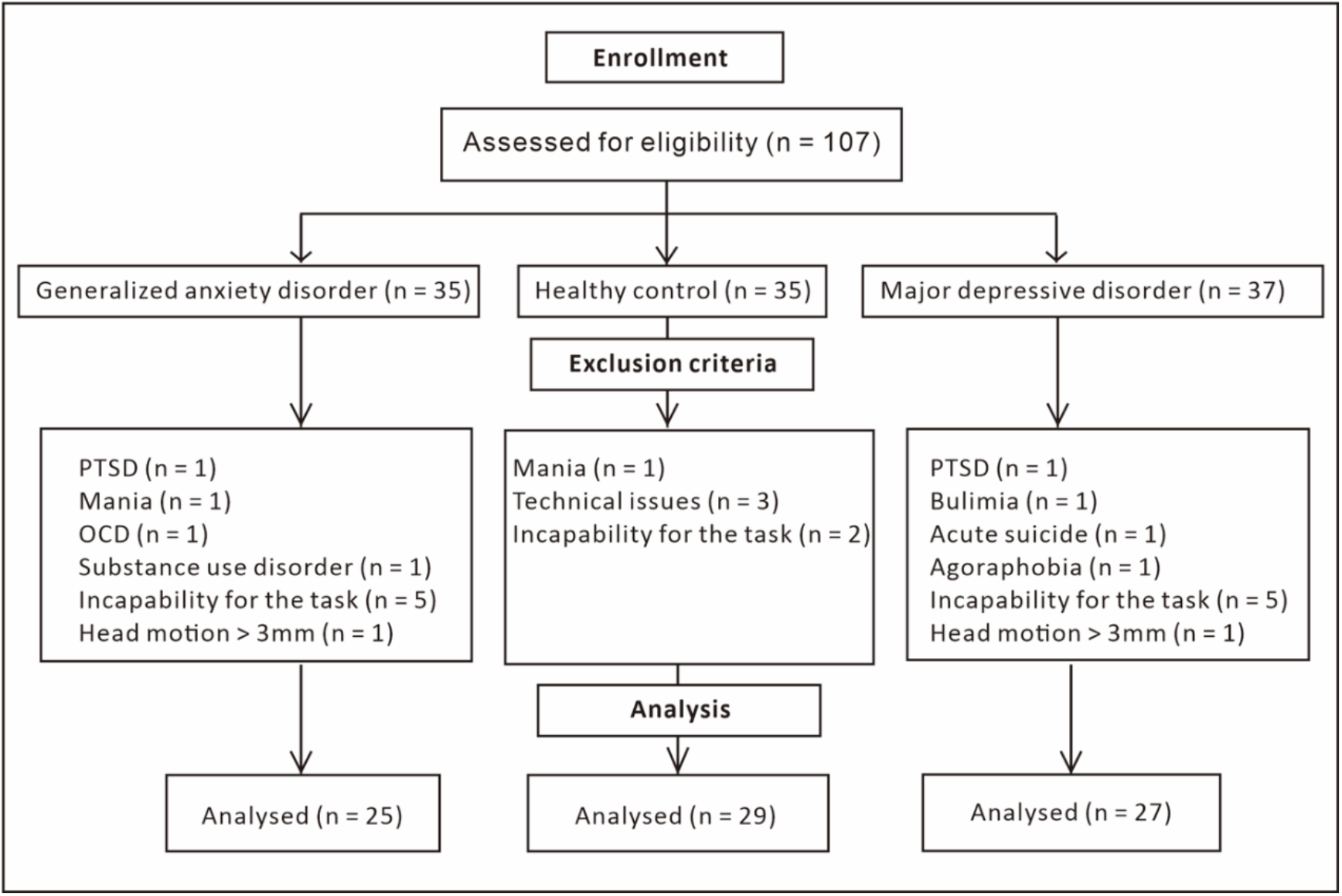
CONSORT FLOW diagram

### MRI data acquisition

MRI data were collected using a 3.0 Tesla GE MR750 system (General Electric Medical System, Milwaukee, WI, USA). Functional images were acquired within a single run using T2*-weighted echo planar imaging (sequence parameters, TR = 2000ms, TE = 30ms, flip angle = 90°, acquisition matrix = 64 × 64, thickness = 3.4mm, FOV = 240 × 240 mm, gap = 0.6mm, slices = 39). High-resolution whole-brain T1-weighted images were additionally obtained to exclude subjects with apparent brain pathologies and to improve normalization of the functional time series (sequence parameters, TR = 6ms, TE = minimum, flip angle = 9°, acquisition matrix = 256 × 256, thickness = 1mm, FOV = 256 × 256mm, slices = 156).

### fMRI data analysis – dimensional analysis

To further account for subclinical co-morbid symptom load the categorical approach (comparing MDD, GAD and HC) was flanked by a dimensional analysis strategy examining associations with MDD and GAD symptom load in the entire sample (pooling the data from MDD, GAD, and HC). In each case the influence of the other symptom dimension was controlled using FSL PALM-alpha110 toolbox (https://fsl.fmrib.ox.ac.uk/fsl/fslwiki/PALM, Permutation Analysis of Linear Models, number of permutations = 5,000) including anxiety or depression symptom load as covariate in the analysis respectively. To assess the collinearity between anxiety and depression symptom load the variance inflation factor (VIF) was calculated. The VIF = 2.37 in this study indicated no problematic collinearity [1-3].

### Behavioral results

The mixed ANOVA load (0-back/1-back) x social (emotion/identity) x disorder (GAD/MDD/HC) with gender as covariate was employed to examine working memory performance in terms of accuracy. The load x social interaction was driven by social condition, such that accuracy was higher for identity recognition across both working memory load conditions (0-back: emotion = 90.5%±0.01, identity = 93.7%±0.01, *p*<0.01; 1-back: emotion = 88.9%±0.01, identity = 92.9%±0.01, *p*<0.01). Disentangling the three-way interaction suggested within-group differences which indicated that MDD patients showed higher accuracy for identity than emotion during 1-back (emotion = 86.7%±0.02, identity = 93.1%±0.02, *p*<0.01) whereas HCs showed higher accuracy for identity than emotion during 0-back (emotion = 87.7% ±0.02, identity = 93.9%±0.02, *p*<0.01).

## Notes

### Competing Interest Statement

The authors have declared no competing interest.

### Author Declarations

Ethics committee of the University of Electronic Science and Technology of China gave ethical approval for this work.

